# Does Internal Preoccupation with Obsessive-Compulsive Themes Affect Externally Oriented Functioning in OCD?: Behavioral Results and Clinical Cases

**DOI:** 10.1101/2022.09.20.22279936

**Authors:** Lora Bednarek, Stephanie Glover, Xiao Ma, Christopher Pittenger, Helen Pushkarskaya

**Affiliations:** Yale School of Medicine; PGSP-Stanford Psy.D. Consortium and Palo Alto University; New York University

**Keywords:** Executive Functioning, Executive Overload Model, Obsessive-Compulsive Disorder, Revised Attention Network Test, Therapeutic Interventions

## Abstract

Individuals with obsessive-compulsive disorder (OCD) exhibit nonspecific deficits in executive function. Internal preoccupations with obsessive-compulsive themes (OCs) may prevent individuals with OCD from fully engaging in externally oriented tasks, explaining these deficits – an ‘executive overload’ model of OCD.

This study reports data from 43 individuals with OCD and 54 healthy individuals collected using the revised Attention Network Test (ANT-R) that is consistent with predictions of the ‘executive overload’ model. During ANT-R, externally orienting cues enhanced individual readiness to respond to external stimuli (alerting benefits), but alerting benefits were negatively associated with severity of internal preoccupations (e.g., neutralizing and obsessing symptoms). Alerting cues improved efficacy of conflict processing (executive benefits), more in individuals with OCD than in healthy controls. These executive benefits correlated positively with the severity of contamination.

Internal preoccupation with OCs could also contribute to poor engagement with exposure and response prevention (ERP) exercises and, consequently, might explain the limited efficacy of ERP-based interventions in some patients. This study describes two clinical cases to illustrate how personalized externally orienting cues may augment ERP exercises to improve patients’ engagement in therapeutic interventions.

The study concludes with discussion of broader implications of the results and with new hypotheses for future investigations.

**Highlights:**

- Internal preoccupations negatively impact executive function in OCD.
- Externally orienting cues improve readiness to respond to external stimuli in OCD.
- Externally orienting cues improve efficacy of conflict processing in OCD.
- Effects of externally orienting cues vary across obsessive-compulsive themes.
- Efficacy of ERP could be improved by augmenting with externally orienting cues.

## Introduction^1^

Obsessive-compulsive disorder (OCD) affects 1-3% of general population and is a leading cause of morbidity worldwide (Kessler et al., 2012; Ruscio et al., 2010). While a formal diagnosis of OCD only requires the presence of distressing and impairing obsessions and/or compulsions, many patients also exhibit difficulties with executive functioning (Shin et al., 2014). These difficulties have been linked to reduced quality of life in affected individuals (Berna Binnur et al., 2005; Subramaniam et al., 2013).

Evidence-based treatments are available, but their efficacy remains limited (Pittenger & Bloch, 2014). Cognitive behavioral therapy (CBT) with exposure and response prevention (ERP) is considered to be highly effective when tailored to an individual’s unique presentation (Reid et al., 2021). It is thought to act through fear reduction and safety learning (McGuire et al., 2014). Estimates of response rates to ERP among OCD patients range from 51% to 97% (Kozak & Foa, 1996; Reid et al., 2021). OCD is markedly heterogeneous (Lochner & Stein, 2003), and some evidence suggested that ERP efficacy may vary across OCD symptom dimensions. Individuals with obsessive-compulsive themes (OCs) related to sexual, religious, unacceptable/taboo, and aggressive thoughts and mental compulsions (internally oriented OCs, *int-OCs*) have been reported to have poorer ERP outcomes (Mataix-Cols et al., 2002; Rufer et al., 2014; Williams et al., 2014), while contamination symptoms (*cont-OCs*) may predict better outcomes (Buchanan et al., 1996). This literature remains mixed (Abramowitz et al., 2003) but suggests that individuals with *int-OCs* may require augmenting ERP-based therapies with additional cognitive interventions (Ferrando & Selai, 2021).

### Difficulties with Executive Functioning in OCD

OCD is associated with broad deficits in multiple domains of executive function – memory, attention shifting, conflict resolution, decision making, and response inhibition – although empirical findings remain inconsistent (Greisberg & McKay, 2003; Kashyap et al., 2013; Shin et al., 2014; Snyder et al., 2015). Neurobiological studies have linked these executive difficulties to altered functioning in cortico-striatal circuits (Nakao et al., 2014); abnormalities in these circuits are also associated with symptoms in OCD (Brennan & Rauch, 2017; Graybiel & Rauch, 2000; Maia et al., 2008).

An executive overload model of OCD (Abramovitch et al., 2012) conceptualizes neuropsychological impairments in OCD as a consequence of ongoing attempts to control the automatic processes associated with obsessive thoughts. Effectively, it suggests that individuals with OCD may be too distracted to attend to the task at hand. This executive overload model of OCD is well aligned with predictions of the neurobiological Default Mode Interference hypothesis, DMN-ih (Sonuga-Barke & Castellanos, 2007). DMN-ih focuses on the dynamic interactions between three large scale brain networks: the default mode network (DMN), the central executive network (CEN), and the salience network (SN) (Bressler & Menon, 2010; Deco et al., 2011). The DMN is involved in self-referential processing of internal states, and is generally deactivated during externally oriented tasks, which instead recruit the CEN (Alves et al., 2019). The SN modulates the DMN↔CEN switch during the rest-task transition (Goulden et al., 2014). DMN-ih posits that the ability of the SN to control the DMN during externally oriented activities is impaired in some individuals, leading to continuous ongoing self-referential processes, such as internally oriented obsessions or mental compulsions in OCD, that distract them from effectively processing external stimuli (Alves-Pinto et al., 2019). DMN-ih is not specific to OCD; other internally oriented processes, such as rumination in depression, may similarly affect performance on externally oriented tasks.

Both the executive overload model of OCD and DMN-ih imply that if a targeted intervention can help individuals with OCD “snap out” of their overfocus on self-referential processes (i.e., give them a “wake-up call”), then their performance on externally oriented tasks should improve. We test this prediction using the revised Attention Network Test, ANT-R (Fan et al., 2009). Next, we present two clinical vignettes that are consistent with predictions of both models. We conclude with a discussion of broader implications of our results.

### Testable hypotheses

The revised Attention Network Test, ANT-R (Fan et al., 2009), builds upon the Three Networks Attentional System model put forward by Posner and Petersen (1990). This model distinguishes among alerting, orienting, and executive attentional networks. The alerting network supports overall awareness and readiness to receive information and respond; the orienting network is concerned with orienting of attention in space and time; the executive attention involves higher level cognition and supports conflict detection and resolution. The alerting network is the most basic attentional network and enables functioning of the other two systems. Importantly, the contribution of alerting to the efficiency of the other networks is not linear, but rather exhibits diminishing marginal return. Thus, when the executive network is operating inefficiently, an alerting stimulus will enhance executive performance, but when the executive network is functioning optimally, the benefit from an alerting stimulus is minimal (Figure 1).

**Figure 1.**
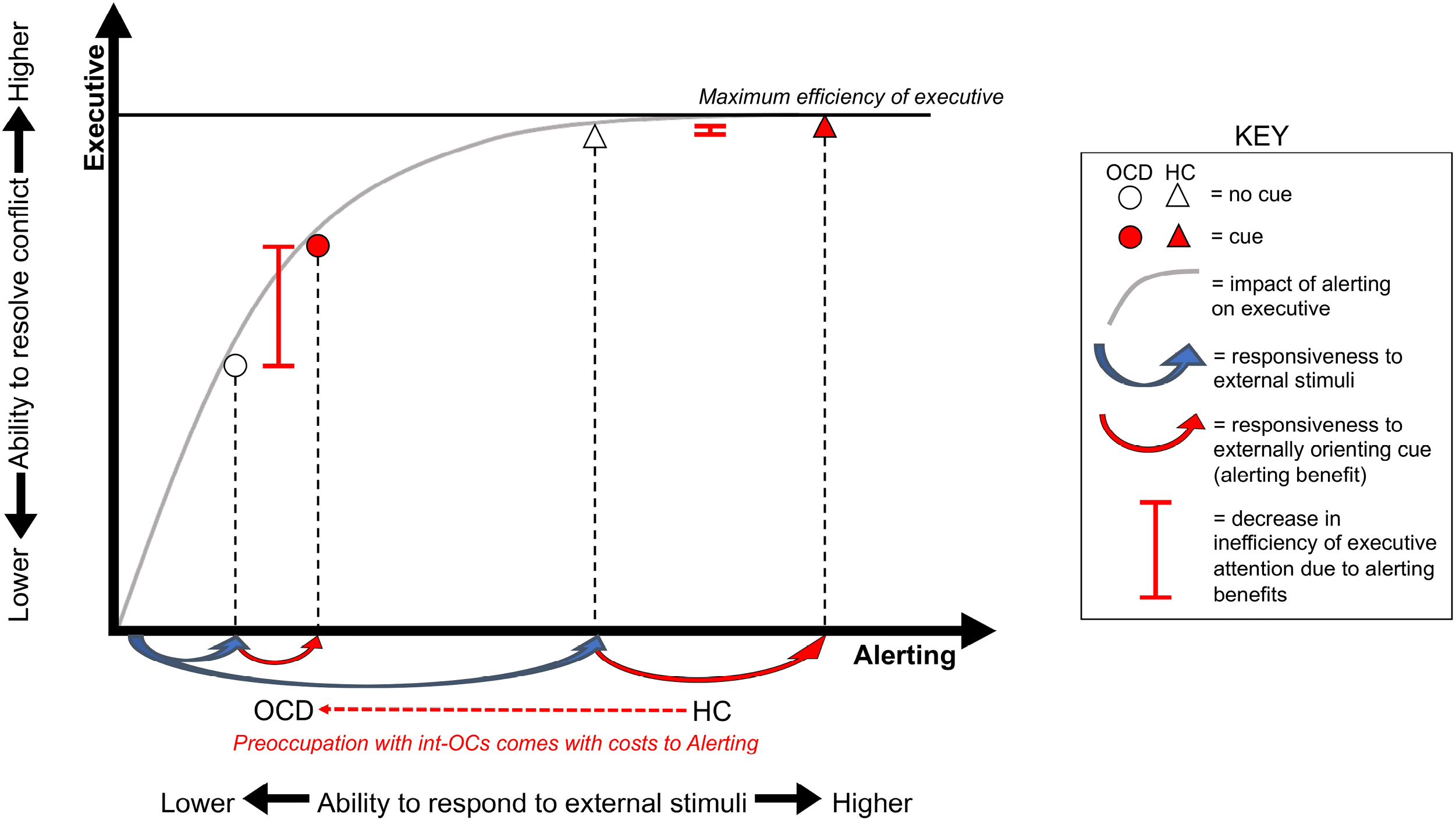
Hypothesized effects of OCs on the efficiency of alerting and executive attention networks. The concave grey line reflects that the positive impact of the efficiency of alerting network on the efficiency of the executive network exhibits diminishing marginal return. The higher efficiency of alerting network is associated with the higher efficiency of executive network, but with the lower additional executive benefits due to the additional alerting benefits. ANT-R (Fan et al., 2009) is calibrated for healthy individuals to be able to perform at a high level of efficiency and, consequently, with very small room for additional improvement in efficiency of the executive network. An executive overload model of OCD (Abramovitch et al., 2012) predicts that internally oriented OCs reduce individual ability to respond to external stimuli (i.e., reduce efficiency of alerting network); this leads to two testable hypotheses. H1: Responsiveness to the external stimuli (target, blue arrows, and cue, red arrows, in ANT-R) is negatively associated with severity of OCs and, thus, is lower in OCD. H2: Efficiency of conflict processing is negatively associated with severity of OCs and, thus, is lower in OCD, and additional executive benefits due to additional alerting benefits are positively associated with severity of OCs and, thus, are higher in OCD.

The ANT-R was designed to probe the efficiency of alerting, orienting, and executive control attention networks, as well as interactions among them, by comparing reaction times (RT, msec) during trials with correct responses, averaged across task conditions. The task was calibrated for participants from non-clinical populations to respond correctly on approximately 80%-90% of trials; faster correct responses are interpreted as indicating higher efficiency of attentional processes (Fan et al., 2009; Mackie et al., 2013).

The ANT-R evaluates how quickly individuals correctly resolve two executive/cognitive control tasks, Simon (Simon & Wolf, 1963) and Flanker (Eriksen & Eriksen, 1974) conflicts. Both tasks require subjects to indicate, with a button press, the direction the middle in a row of five arrows is pointing. On Simon trials, there is conflict between the location of the correct response button and the side of the screen where the stimulus appears (e.g., a left-pointing middle arrow requires to press the button on the left side of the keyboard but appears on the right side of the screen). On Flanker trials, there is conflict between the direction of the middle arrow and that of the adjacent ‘flanking’ arrows.

In the ANT-R, on some trials, participants receive a warning – a “wake-up call” – a few hundred milliseconds before stimulus onset (Figure 2). Participants tend to respond correctly more quickly when they received such warnings; this is interpreted as improvement in alerting due to the externally orienting cue. Furthermore, participants tend to be more efficient in resolving Simon and Flanker conflicts on trials when they receive “wake-up calls” (a positive alerting x executive interaction); this is interpreted as an impact of alerting benefits on the efficiency of executive attention. Note that this effect depends on both the alerting effect and the baseline efficiency of executive attention (see Figure 1).

**Figure 2.**
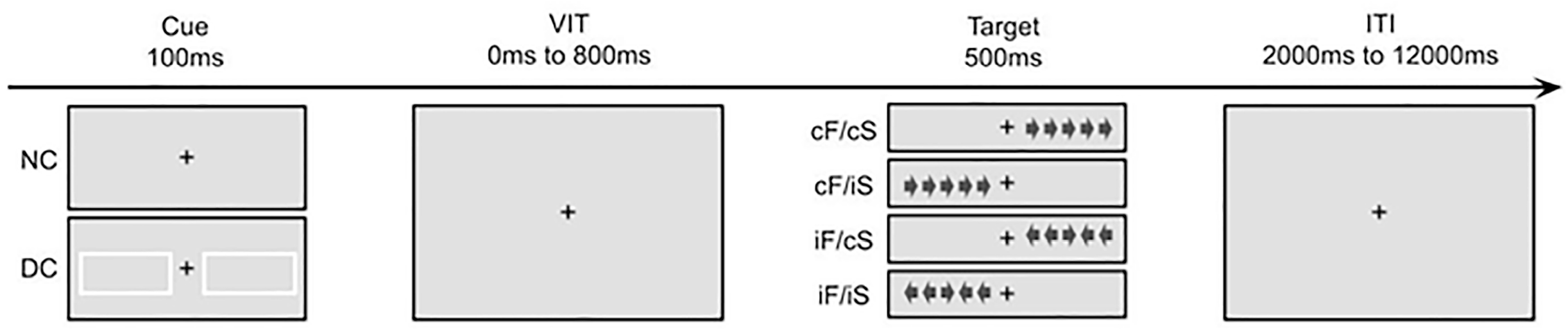
Schematic presentation of components of the revised Attention Network Test (ANT-R, Fan et al., 2009), relevant to study hypotheses. ANT-R consists of 4 sequential phases. Phase I, Cue: in the beginning of some trials, two cue boxes flash for 100ms (double cue, DC); no boxes appear during some other trials (no cue, NC). Phase II, Variable interval, VIT: a fixation cross appears in the middle of the screen for 0, 400, or 800ms. Phase III, Target: five arrows are presented on the left or the right side of the fixation cross for 500 ms. Participants are asked to indicate where the middle arrow (target) points by pressing key “A” (the left side of the keyboard) if it points to the left or key “L” (the right side of the keyboard) if it points to the right. During congruent Simon (cS) trials, the arrows and the correct response button are located on the same side; during incongruent Simon (iS) trials, the arrows and the correct response button are located on opposite sides; during congruent Flanker (cF) trials all five arrows point in the same direction; during incongruent Flanker (iF) trials, the target arrow and the surrounding arrows point in opposite directions. Overall, there are four types of trials: cF/CS, cF/iS, iF/cS, and iF/iS. Phase IV, Inter Trials Interval, ITI: A fixation cross appears in the middle of the screen for 2000, 4000, 6000, 8000, 10000, or 12000 ms (mean = 4000 ms). *Note*. Picture depicts stimuli for the correct response ‘L’, a key located on the right side of the keyboard.

We predict that overfocus on internal processes is associated with a reduced ability to attend to external stimuli. In OCD, such overfocus maybe linked to the severity of OCs, particularly *int-OCs*. In the context of ANT-R, this suggests that both readiness to respond to the target stimuli (Figure 1, blue arrows) and the benefit provided by an externally orienting cue (Figure 1, red arrows) should be negatively associated with severity of OCs. This leads to two sets of testable hypotheses.

First, we anticipate an improvement in RT during correct trials following “wake-up calls” (i.e., alerting benefits) in all study participants, but we anticipate that these improvements should be reduced in OCD, and negatively associated with severity of OCs. Given the significant heterogeneity of OCs, we anticipate that some of the OC dimensions maybe linked to reduced alerting benefits more than others.

> *H1*: Alerting benefits are lower in OCD than in healthy controls.
>
> *H1a:* Association of alerting benefits with severity of OCs varies across OCD symptom dimensions.

Second, recall that ANT-R is calibrated for participants from non-clinical populations to perform at a high level of efficiency during Simon/Flanker conflict processing (see Figure 1). If readiness to respond to target stimuli is negatively associated with severity of OCs, then efficiency of Simon/Flanker conflict processing is also negatively associated with severity of OCs and is expected to be lower in individuals with OCD than in healthy control group. However, alerting benefits (even if they are reduced) may lead to a larger improvement in efficacy of conflict processing in OCD participants than in healthy controls (see Figure 1). Given the significant heterogeneity of OCs, we anticipate that some of the OC dimensions maybe linked to improvement in efficacy of conflict processing more than others.

> *H2*: Alerting benefits result in stronger improvement in efficiency of executive attention in individuals with OCD than in healthy controls.
>
> *H2a:* The impact of alerting benefits on efficiency of executive attention varies across OCD symptom dimensions.

Note that hypotheses H2/H2a depend on the assumption that healthy controls operate close to maximum efficiency during ANT-R (i.e., on the flat portion of the curve in Figure 1); if this assumption is violated, then stronger alerting benefits in healthy controls could result in greater improvements in efficiency of executive attention in healthy controls than in individuals with OCD.

### Potential confounds

We control for effects of two potentially important confounds, symptoms of attention deficit-hyperactivity disorder and depressive symptoms. Both are common comorbidities of OCD (Pallanti et al., 2011) that may contribute to neuropsychological deficits (Brem et al., 2014; Moritz et al., 2001). For instance, studies that used ANT-R reported that ADHD affects alerting and executive attention (Arora et al., 2020; Berger & Posner, 2000) and that depression affects executive attention (Wang et al., 2020).

## Methods

### Participants

All procedures were approved by the Yale University Human Investigation Committee. A total of 97 unmedicated participants were recruited through the Yale OCD Research Clinic between 2012 and 2020 by posting flyers in the local community as a part of a larger series of investigations (Ma et al., 2021; Pushkarskaya et al., 2019; Pushkarskaya et al., 2015). The sample included 43 unmedicated individuals with clinically significantly OCs (OCD) and 54 individuals with no psychiatric diagnosis (HC). All provided written informed consent before any procedure.

Diagnoses were established by the Mini International Neuropsychiatric Interview and confirmed by doctoral-level clinicians. Clinically significant OCs were defined as 16 or higher on the Yale-Brown Obsessive Compulsive Scale (Y-BOCS; Goodman et al., 1989a,b). OCD was the primary clinical diagnosis in all patients; comorbidities included depression, hoarding disorder, panic disorder, social phobia, agoraphobia, post-traumatic stress disorder, and generalized anxiety disorder.

### Procedure

Participants completed a demographic questionnaire, the Kaufman Brief Intelligence Test (Kaufman & Kaufman, 2004), the revised Obsessive-Compulsive Inventory (OCI-R; (Foa et al., 1998), the Beck Depression Inventory II (BDI-II; Beck et al., 1996), the Adult ADHD Self-Report Scale (ASRS; Adler et al., 2006), and the revised Attention Network Test (ANT-R). They were compensated for their time.

### Measures

#### Clinical measures

The OCI-R is an 18-item 5-point Likert-type self-report scale of how much obsessive-compulsive symptoms have been distressing or bothersome them during the past month (from 0 = “not at all” to 4 = “extremely”). The OCI-R has been shown to have good internal consistency, convergent validity, and test-retest reliability, in both clinical and nonclinical samples (Foa et al., 1998). The OCI-R includes six subscales (3 items each) that have been shown to map well onto items from Y-BOCS symptom checklist (Huppert et al., 2007): washing, obsessing, hoarding, checking, ordering, and neutralizing. Higher scores along obsessing and neutralizing dimensions reflect stronger *int-OCs* tendencies. *Obsessing* denotes intrusive thoughts of harm to self or others, including taboo, sexual, or aggressive thoughts (e.g., *“I frequently get nasty thoughts and have difficulty in getting rid of them”*). *Neutralizing* reflects mental compulsions, such as counting items or actions (e.g., *“I feel compelled to count while I am doing things”*).

The BDI-II is a 21-item 4-point self-report scale designed to measure the severity of depressive symptomatology during the past month. The BDI-II is widely used as an indicator of the severity of depression, and numerous studies provide evidence for its reliability and validity across different populations and cultural groups (Wang & Gorenstein, 2013).

The ASRS is an 18-item self-report questionnaire designed to assess attention deficit-hyperactivity disorder (ADHD) symptoms in adults. Part A, consisting of 6 items, has been found to be the most predictive of ADHD and are best for use as a screening instrument. Part B consists of 12 additional questions that provide additional cues and can serve as further probes into the symptoms. The ASRS has high internal consistency and concurrent validity (Adler et al., 2006).

### The Revised Attention Network Test

The ANT-R is described in detail in Fan et al. (2009). Participants are presented with five arrows (the stimulus) on the right or the left side of the screen and are asked to press a button (on the left or on the right side of the keyboard) corresponding to the direction of the middle arrow (the target; see Figure 2). Shortly before the stimulus appears, a cue (two lighted square frames) is presented during some trials (double cue), while during some trials no cue is presented (no cue). There are also trials on which a single pre-stimulus cue is presented, on the left or right side of the screen, but they are not analyzed here. The button for the correct response is located on the same side as the stimulus on some trials (congruent Simon) and on the opposite side on other trials (incongruent Simon). The target arrow points in the same direction as the remaining 4 arrows on some trials (congruent Flanker) and in the opposite direction on others (incongruent Flanker). Participants tend to respond correctly to incongruent conditions (direction/location) more slowly than to congruent conditions (Flanker/Simon; Hübner and Töbel, 2019), since the former require more executive attention effort.

Participants practiced the task for three minutes. After they indicated they understood the task, they were invited to begin the experiment.

### Behavior-based measures

ANT-R measures are computed based on reaction time (RT, ms) on trials on which participants responded correctly.

*Alerting benefits* (“*Alerting*”) are measured as:

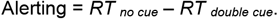

A higher alerting value indicates greater improvement in performance on the task following a “wake-up call” (i.e., faster correct responses).

*Executive benefits (“Executive”)* are quantified by two measures:

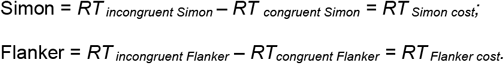

A higher executive value indicates higher costs of Simon/Flanker conflict processing and implies greater inefficiency of executive attention. In this paper we do not analyze Simon/Flanker interaction.

*Effects of Alerting on Executive Attention (“Alerting x Executive”)* are quantified by two measures:

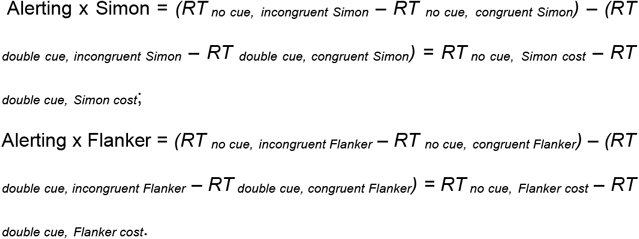

More positive values indicate stronger improvement in efficiency of conflict processing (Simon or Flanker) following a “wake-up call” and imply higher baseline degree of inefficiency of executive attention.

### Analyses

To test H1 and H2, while controlling for potential confounds, we used a subset of our participants who (i) completed ANT-R, demographic questionnaire, Kbit, BDI-II, and ASRS, and (ii) allowed us to match OCD and HC on age, gender, and IQ (*matched subsample*).

To test H1a and H2a, while controlling for potential confounds, we used a subset of our participants who completed ANT-R, demographic questionnaire, Kbit, BDI-II, ASRS, and OCI-R (*OCI-R subsample*). We ran a series of linear regression analyses with OCI-R subscales as dependent variables and IQ, age, BDI-II, and ASRS as covariates. The standardized residuals from these regressions (OCI-R subscales stdres) strongly correlated with respective OCI-R subscales but did not significantly correlate with potentially confounding variables (Appendix B). Thus, associations of these standardized residuals with ANT-R measures are independent of the potential associations between IQ, age, BDI-II, and ASRS and ANT-R measures. Appendix E reports results from analyses that do not control for these potential confounding effects.

To test H1 and H1a, we used a series of univariate ANCOVAs with alerting as the dependent variable. For H1, the between-subject factor was diagnosis (OCD/HC), and IQ, age, BDI-II, and ASRS were covariates. For H1a, OCI-R stdres subscales (washing, obsessing, hoarding, ordering, checking, and neutralizing), IQ, age, BDI-II, and ASRS were covariates. All covariates were standardized.

To test H2 and H2a, we used a series of repeated measures ANCOVAs with *Alerting x Executive* as the dependent variable and type of conflict (Simon/Flanker) as the repeated measure. For H2, the between-subject factor was diagnosis (OCD/HC), and IQ, age, BDI-II, ASRS, and alerting were covariates. For H2a, OCI-R subscales stdres (washing, obsessing, hoarding, ordering, checking, and neutralizing), IQ, age, BDI-II, ASRS, and alerting were covariates. All covariates were standardized.

To ensure that assumptions of ANOVA were not violated, we examined the residuals from each ANCOVA for (near) normality (using the Shapiro Wilks test and evaluating skewness) and for the presence of outliers (using z-scores ≥ |3| criteria).

## Results

### Sample and subsamples

97 unmedicated participants (43 OCD, 54 HC) completed ANT-R and the demographic questionnaire; 96 of them additionally completed Kbit; 87 of them additionally completed BDI-II and ASRS; 84 of them additionally completed OCI-R (Appendix A). Two subsamples of these participants were included in two sets of analyses, as detailed below.

35 OCD and 52 HC completed ANT-R, the demographic questionnaire, Kbit, BDI-II, and ASRS. Reliabilities for BDI-II and ASRS were excellent (Cronbach’s α = 0.94 and α = 0.87, respectively). From these individuals, we selected a subset of 35 OCD (12 males) and 46 HC (23 males) to match the two diagnostic groups on age, gender, and IQ (*matched subsample*, to test H1 and H2; Table 1).

**Table 1.** Demographic characteristics of study participants.

|  | All<br>participants |  | OCD |  | HC | test<br>stats | p |
| --- | --- | --- | --- | --- | --- | --- | --- |
|  | mean (SD) | range | mean (SD) | range | mean (SD) |  |  |
| *Matched subsample, N = 81* |  |  | N = 35 |  | N = 46 |  |  |
|  |  |  |  |  |  | X<sup>2</sup> |  |
| Male, % | 31 (38) |  | 12 (34) |  | 19 (41) | 0.41 | 0.52 |
|  |  |  |  |  |  | ANOVA, F-stat |  |
| Age | 32.7 (11.8) | 18 - 62 | 34.2 (12.3) | 18 - 59 | 31.5 (11.4) | 1.0 | 0.32 |
| IQ | 109 (13) | 84 - 130 | 106 (13) | 77 - 130 | 111 (13) | 2.9 | 0.09 |
|  |  |  |  |  |  | Mann Whitney U |  |
| BDI | 6.8 (8.9) | 0 - 36 | 11.8 (10.5) | 0 - 20 | 3.0 (4.7) | 1226 | <0.001 |
| ASRS | 5.1 (4.7) | 0 - 18 | 6.5 (5.7) | 0 - 13 | 4.0 (3.8) | 1001 | 0.06 |
| *OCI-R subsample, N = 83* |  |  | N = 32 |  | N = 51 |  |  |
|  |  |  |  |  |  | X<sup>2</sup> |  |
| Male, n (%) | 35 (42) |  | 12 (37) |  | 23 (45) | 0.47 | 0.50 |
|  |  |  |  |  |  | ANOVA, F-stat |  |
| Age | 31.9 (11.8) | 18 - 62 | 33.9 (12.6) | 18 - 59 | 30.7 (11.2) | 1.5 | 0.23 |
| IQ | 111 (14) | 84 - 130 | 107 (12) | 77 - 132 | 113 (14) | 4.5 | 0.04 |
|  |  |  |  |  |  | Mann Whitney U |  |
| BDI | 6.5 (8.7) | 0 - 36 | 11.7 (10.8) | 0 - 20 | 3.2 (4.8) | 1204 | <0.001 |
| ASRS | 5.0 (4.8) | 0 - 18 | 6.3 (5.9) | 0 - 13 | 4.2 (3.8) | 949 | 0.21 |
| Washing | 2.4 (3.5) | 0 -12 | 5.4 (4.0) | 0 - 3 | 0.6 (1.0) | 1433 | <0.001 |
| Obsessing | 1.6 (2.5) | 0 - 10 | 3.4 (3.0) | 0 - 4 | 0.9 (0.1) | 1298 | <0.001 |
| Hoarding | 1.7 (2.1) | 0 - 11 | 2.4 (2.7) | 0 - 7 | 1.3 (1.6) | 1016 | 0.05 |
| Ordering | 3.5 (3.5) | 0 - 12 | 6.0 (3.9) | 0 - 7 | 1.8 (2.0) | 1353 | <0.001 |
| Checking | 2.4 (3.1 ) | 0 - 11 | 4.5 (3.3) | 0 - 6 | 0.8 (1.5) | 1438 | <0.001 |
| Neutralizing | 1.4 (2.5) | 0 - 12 | 2.8 (3.5) | 0 - 5 | 0.5 (1.1) | 1142 | <0.001 |

32 OCD and 51 HC completed ANT-R, the demographic questionnaire, Kbit, and all three clinical scales of interest (*OCI-R subsample*, to test H1a and H2a; Table 1). Reliability for OCI-R was excellent (Cronbach’s α = 0.92), and for OCI-R subscales ranged from good (Cronbach’s α = 0.75, hoarding) to excellent (Cronbach’s α = 0.93, for washing). OCI-R scores were skewed toward 0 (Shapiro Wilks ps <0.001, skewness >1.03), since many healthy individuals self-reported minimal to no OCs. On average, individuals with OCD scored higher than did HC (OCD: mean 24.84 ± 11.73; HC: mean 5.35 ± 6.00; Mann-Whitney U p < 0.001); but OCI-R distributions were broad and overlapping across diagnostic groups (Appendix A). Some individuals with OCD self-reported relatively low scores on OCI-R (OCI-R < 10), despite clinically significant OCs (Y-BOCS ≥ 16, see *Participants*), which could indicate a tendency to minimize their symptoms.

As expected, BDI-II and ASRS scores significantly correlated with OCI-R total scale and subscales, but not with their standardized residuals (Appendix B). Most OCI-R subscales also correlated, as did their standardized residuals (Appendix B), but these relations were not strong enough to raise multicollinearity concerns (Spearman’s ρ ≤ 0.60, Dormann et al. (2013)).

### H1, H1a: Alerting Benefit and severity of OCs

In the full sample, alerting benefits were positive (except for 3 OCD and 2 HC) and normally distributed (Shapiro Wilks p = 0.29).

As predicted (Figure 1), in the *matched subsample*, OCD tended to respond qualitatively more slowly than HC (median for OCD: RT_no cue_ = 676±117SD, RT_double cue_ = 626±106SD; median for HC: RT_no cue_ = 651±108SD & RT_double cue_ = 618±97SD), with accuracy being high and comparable across two groups (median for OCD = 0.97±0.13SD, median for HC = 0.96±0.07SD; Mann-Whitney U p = 0.36). RT improvement from the no cue condition to the double cue condition was qualitatively smaller in OCD than in HC (mean for OCD = 38±30SD; mean for HC = 46±33SD). For details see Appendix C. However, there was significant within-group variability (see above) and, in contrast to H1, univariate ANCOVA in the matched subsample did not reveal a significant effect of OCD diagnosis on alerting benefits (Table 2). The residuals from this ANCOVA were normally distributed (Shapiro-Wilk p = 0.57).

**Table 2.** Testing H1: univariate ANCOVA with dependent variable Alerting.

| Predictor | effect |  | ANCOVA |  | parameter estimates |  |
| --- | --- | --- | --- | --- | --- | --- |
| | direction | size, $\eta^2$ | F | p | $\beta$ | SE |
| Constant | + | 1.00 | 518.60 | 0.49 | 38.89 | 6.07 |
| age, std | + | <0.001 | 0.04 | 0.85 | 0.71 | 3.75 |
| IQ, std | - | 0.002 | 0.17 | 0.68 | -1.56 | 3.77 |
| ASRS, std | - | 0.004 | 0.32 | 0.57 | -2.57 | 4.55 |
| BDI, std | - | 0.001 | 0.05 | 0.82 | -1.13 | 5.01 |
| Dx | + | 0.01 | 0.47 | 0.50 | 5.89 | 8.62 |

*Note:* Effect directions + - positive, - - negative. Effect sizes $\eta^2 = 0.01$ - small effect, $\eta^2 = 0.06$ - medium effect, $\eta^2 = 0.14$ - large effect. Covariates std – standardized.

Consistent with H1a, univariate ANCOVA in the *OCI-R subsample* (Table 3) revealed that, when we control for potential confounds, alerting benefits were negatively associated with neutralizing (neutralizing stdres: β = -8.77, F = 4.28 p = 0.04, η^2^ = 0.06 medium effect size); we also observed a trend toward negative association of alerting with obsessing (obsessing stdres β = -8.18, F = 2.79 p = 0.10, η^2^ = 0.04 small-to-medium effect size) and positive association of alerting with ordering (ordering stdres β = 10.90, F = 3.93 p = 0.052, η^2^ = 0.05 small-to-medium effect size). The residuals from this ANCOVA were near normally distributed (Shapiro-Wilk p = 0.03, skewness = 0.37, no extreme outliers).

**Table 3.** Testing H1a: univariate ANCOVA with dependent variable Alerting.

| Predictor | effect |  | ANCOVA |  | parameter estimates |  |
| --- | --- | --- | --- | --- | --- | --- |
| | direction | size, $\eta^2$ | F | p | $\beta$ | SE |
| Constant | + | 0.03 | 2.36 | 0.13 | 49.52 | 32.24 |
| age, std | + | 0.003 | 0.20 | 0.66 | 0.13 | 0.30 |
| IQ, std | - | 0.002 | 0.17 | 0.68 | -0.11 | 0.26 |
| ASRS, std | - | 0.010 | 0.76 | 0.39 | -3.88 | 4.45 |
| BDI, std | - | 0.006 | 0.42 | 0.52 | -2.79 | 4.29 |
| Washing, stdres | + | 0.01 | 1.04 | 0.31 | 4.88 | 4.78 |
| Obsessing, stdres | - | 0.04<sup>a</sup> | 2.79 | 0.10 | -8.18 | 4.90 |
| Hoarding, stdres | - | 0.02 | 1.55 | 0.22 | -5.22 | 4.19 |
| Ordering, stdres | + | 0.05<sup>*</sup> | 3.93 | 0.05 | 10.90 | 5.50 |
| Checking, stdres | - | 0.004 | 0.26 | 0.61 | -2.85 | 5.57 |
| Neutralizing, stdres | - | 0.06<sup>*</sup> | 4.28 | 0.04 | -8.77 | 4.24 |

*Note:* Significance levels \* - $p < 0.05$ level; <sup>a</sup> - $p < 0.1$ . Effect directions + - positive, - - negative. Effect sizes $\eta^2 = 0.01$ - small effect, $\eta^2 = 0.06$ - medium effect, $\eta^2 = 0.14$ - large effect. Covariates std – standardized, stdres – standardized residuals.

None of the control covariates (age, IQ, BDI, and ASRS) significantly related to alerting in either ANCOVA.

Overall, these results provide only trend-level support for H1 but are consistent with H1a and with our expectations that alerting benefits are negatively associated with severity of *int-OCs*. Analyzing OCD participants only (excluding controls) revealed similar patterns (Appendix C).

### H2, H2a: Impact of Alerting Benefit on Executive Attention and severity of OCs

In the full sample, on average, RT costs to successfully process Flanker conflict were higher than RT costs to successfully process Simon conflict, suggesting that Flanker is a more difficult cognitive control task. OCD took longer than HC to process both conflicts during no cue trials (OCD: RT_no cue, Simon cost_ = 18 msec, RT_no cue, Flanker cost_ = 163 msec; HC: RT_no cue, Simon cost_ = 8 msec, RT_no cue, Flanker cost_ = 153 msec); OCD performance from no cue trials to double cue trials improved more than that of HC (OCD: Alerting x Simon = 20 msec, Alerting x Flanker: 12 msec; HC: Alerting x Simon = 4 msec, Alerting x Flanker: 2 msec).

In the *matched subsample*, repeated measures ANCOVA revealed that the effect of alerting on executive attention was significantly stronger in OCD (main effect of OCD: F = 12.24 p < 0.001, η^2^ = 0.14, large effect size; Figure 3, Table 4). This effect did not differ significantly between Simon and Flanker processing (OCD x conflict type: F = 0.81, p = 0.37, η^2^ = 0.01, small effect size), suggesting that improved readiness to respond led to broad improvement in executive attention in individuals with OCD (H2).

**Table 4.** Testing H2: repeated measures ANCOVA with dependent variable Alerting x Executive, and repeated measures type of conflict (Simon/Flanker)

| Predictors | Main effects |  |  |  | Predictor x conflict interactions |  |  |  |
| --- | --- | --- | --- | --- | --- | --- | --- | --- |
|  | effect | ANCOVA |  |  | effect | ANCOVA |  |  |
| | direction | size, $\eta^2$ | F | p | direction | size, $\eta^2$ | F | p |
| Intercept | + | 0.13 | 10.72 | 0.00 | S > F | 0.007 | 0.51 | 0.48 |
| age, std | - | 0.02 | 1.17 | 0.28 | S < F | 0.001 | 0.08 | 0.78 |
| IQ, std | + | 0.02 | 1.69 | 0.20 | S > F | 0.003 | 0.21 | 0.65 |
| ASRS, std | - | 0.05<sup>a</sup> | 3.77 | 0.06 | S > F | 0.002 | 0.16 | 0.69 |
| BDI, std | + | <0.001 | 0.001 | 0.98 | S > F | <0.001 | <0.001 | 0.99 |
| Alerting, std | + | 0.08* | 6.26 | 0.02 | S < F | 0.09** | 7.06 | 0.01 |
| Dx | + | 0.14*** | 12.24 | <.001 | S > F | 0.01 | 0.81 | 0.37 |

*Note:* Significance levels \*\*\* - $p < 0.001$ ; \*\* - $p < 0.01$ ; \* - $p < 0.05$ level; <sup>a</sup> - $p < 0.1$ . Main effect directions + - positive, - - negative; difference of effects across types of conflicts S>F – effect is stronger under Simon conflict, S < F – effect is stronger under Flanker conflict. Effect sizes $\eta^2 = 0.01$ - small effect, $\eta^2 = 0.06$ - medium effect, $\eta^2 = 0.14$ - large effect. Covariates std – standardized.

**Figure 3.**
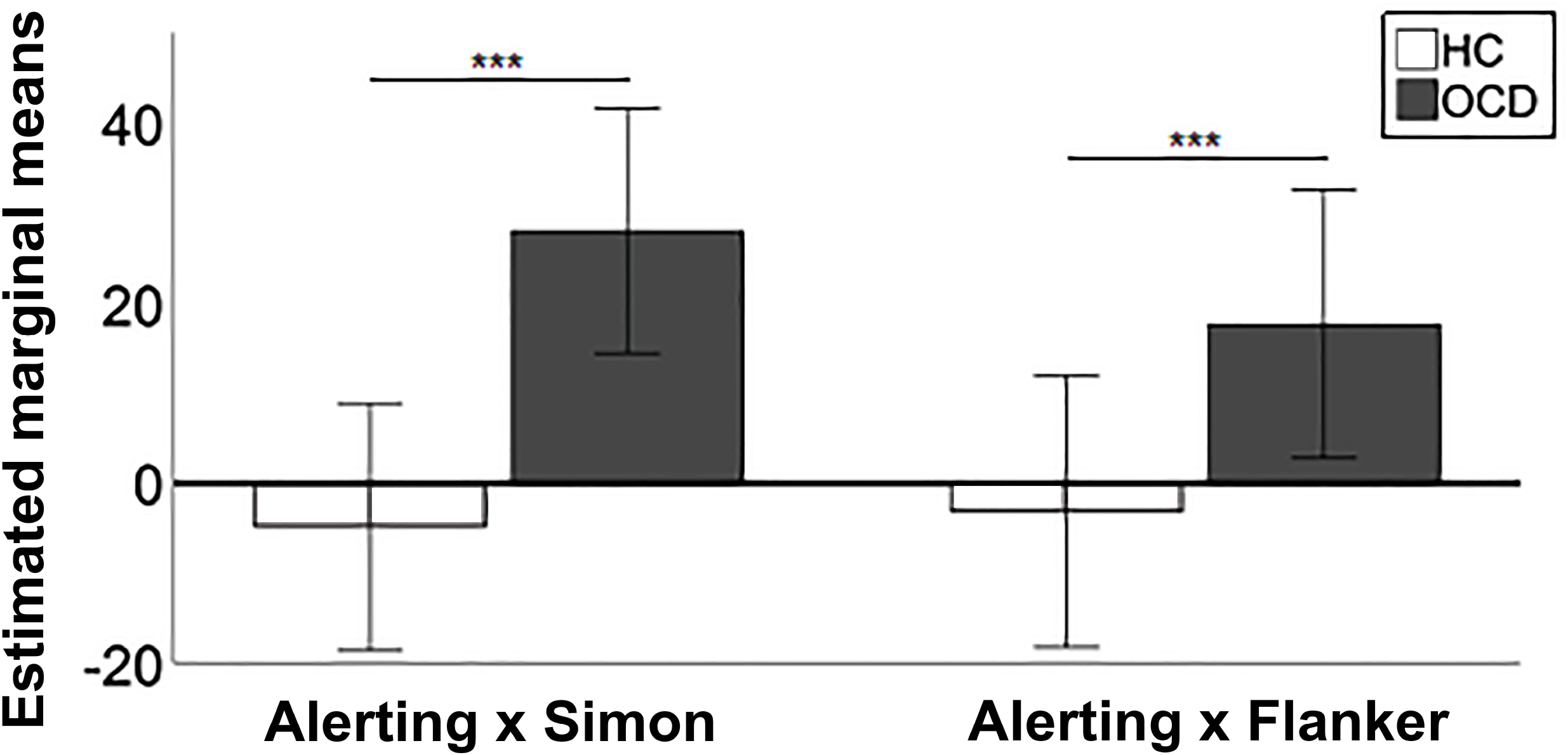
Effects of improvement in readiness to respond due to externally orienting cues (i.e., Alerting benefits) on efficacy of Simon (Alerting x Simon) and Flanker (Alerting x Flanker) conflict processing. Estimated marginal means from Repeated Measures ANCOVA conducted on data from *the matched subsample* (43 individuals with OCD and 43 healthy individuals); see Methods for details. *Note:* OCD = individuals with Obsessive-compulsive disorder, HC = healthy individuals. Significance levels *** - p < 0.001; Error bars 95% Cl.

Individual responsiveness to externally orienting cues (i.e., alerting benefit) was positively associated with improvement in Flanker conflict processing (β = 17.18, t = 3.22 p = 0.002, η^2^ = 0.12, medium-to-large effect size) but not with improvement in Simon conflict processing (t = 0.25 p = 0.80, η^2^ = 0.001; alerting x conflict type interaction F = 7.06 p = 0.01). This is consistent with our earlier observations that Flanker maybe a more difficult conflict to process: our participants are likely to operate at a lower level of efficiency, and, thus, changes in effectiveness of Flanker processing are more sensitive to changes in alerting level than in Simon processing (see Figure 1).

Effects of control variables, age, IQ, and severity of depressive symptoms did not have significant effects in this subsample. Severity of ADHD symptoms negatively correlated with alerting x executive at trend level (F = 3.77 p = 0.06, η^2^ = 0.05 small-to-medium effect size), suggesting that impact of alerting on executive attention may be reduced in individuals with more severe ADHD symptoms. The residuals from this ANCOVA were normally distributed (Alerting x Simon: Shapiro-Wilk p = 0.84, Alerting x Flanker: Shapiro-Wilk p = 0.62).

In the *OCI-R subsample*, severity of washing symptoms positively correlated with the effect of alerting in both Simon and Flanker conflicts (washing stdres: main effect F = 8.86 p = 0.004, η^2^ = 0.12, medium-to-large effect size; washing x conflict interaction: F = 0.26, p = 0.61, η^2^ = 0.004; Table 5).

**Table 5.**
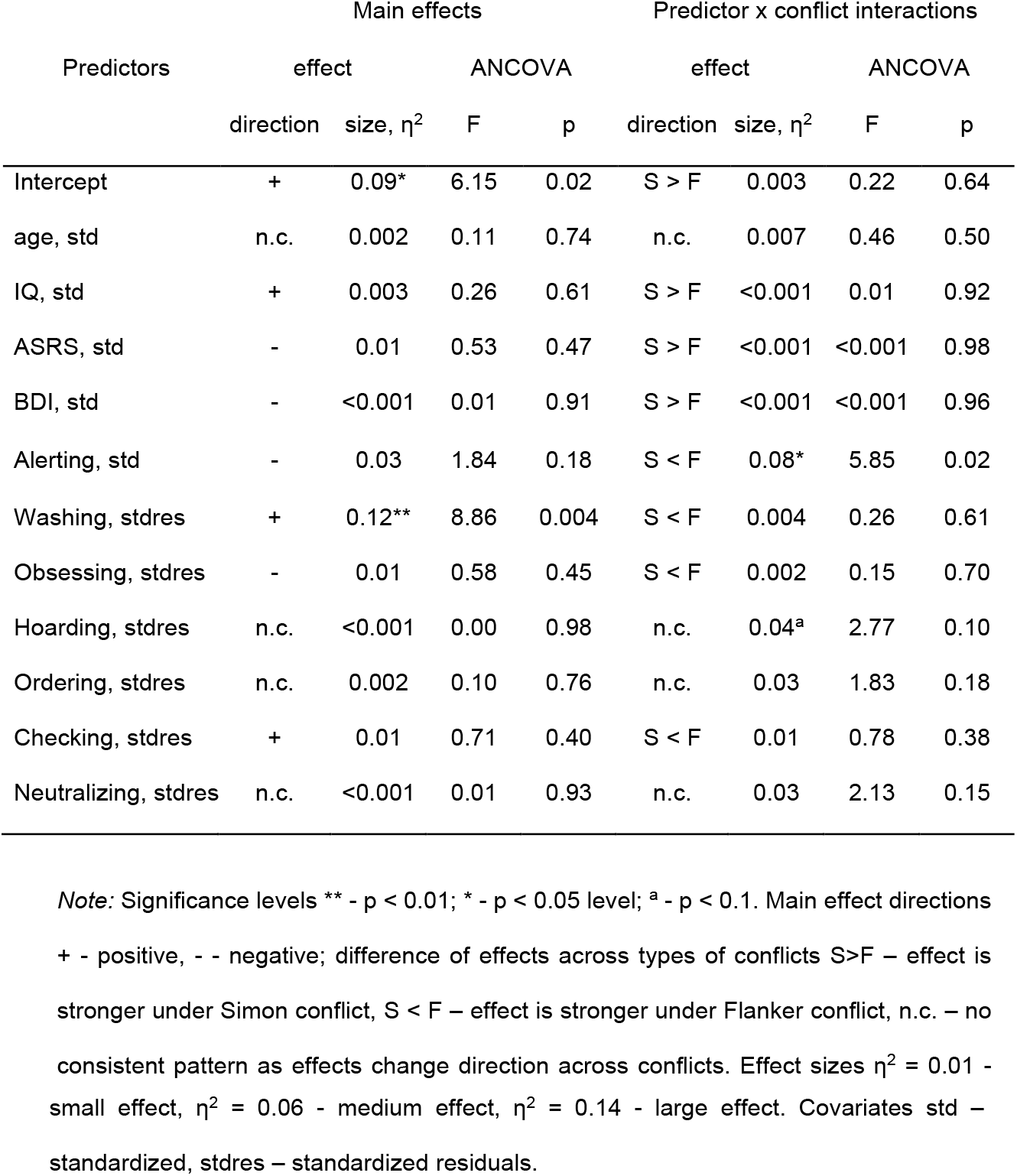
Testing H2a: repeated measures ANCOVA with dependent variable Alerting x Executive, and repeated measures type of conflict (Simon/Flanker)

As in the matched subsample, individual responsiveness to externally orienting cues (i.e., alerting benefit) was positively associated with improvement in efficiency of Flanker conflict processing (β = 15.09, t = 2.50 p = 0.015, η^2^ = 0.08, medium-to-large effect size) but not with improvement in Simon conflict processing (t = -1.02 p = 0.31, η^2^ = 0.014 small effect size; alerting x conflict interaction F = 5.85 p = 0.02). Effects of control variables, age, IQ, and severity of depressive and ADHD symptoms did not have significant effect in this subsample.

The residuals from this ANCOVA we were (near)normally distributed (Alerting x Simon: Shapiro-Wilk p = 0.50, Alerting x Flanker: Shapiro-Wilk p = 0.07, no extreme outliers).

In exploratory analyses, we examined whether gender modulates any relations of interest in univariate and repeated measures ANCOVAs detailed above. None of the significant gender effects were detected (ps > 0.10).

### Summary of the behavioral results

Overall, these behavioral results are consistent with our hypotheses and indicate that individuals with OCD tend to improve their performance on cognitive control tasks (Simon and Flanker) when they receive an externally orienting cue (a “wake-up call”). Individuals with more severe *int-OCs* (neutralizing and obsessing symptoms) have more difficulty “snapping out” from the internal preoccupation when prompted to attend to external stimuli. In contrast, individuals with more severe *cont-OCs* appear to be able to benefit the most from an externally orienting cue during cognitive control tasks.

## Clinical Relevance of Behavioral Results

A recent meta-analysis suggested that ERP-based therapies may benefit from augmentation with additional cognitive interventions (Ferrando & Selai, 2021), such as mindfulness (Wilkinson-Tough et al., 2010), autogenic (Nakatani et al., 2005), or relaxation trainings (Fineberg et al., 2005). To add to this literature, our results indicate that externally orienting cues (“wake-up calls”) may help patients with OCD break away from their obsessive thoughts, re-orient to the present moment and improve their cognitive control abilities in the context of an ongoing exposure and response prevention exercise. Using subtle external cues may be enough to help individuals with *cont-OCs* re-orient to tasks at hand, whereas more active, compelling cues may be required in individuals with *int-OCs*. The following clinical vignettes (each is cumulatively based on a variety of cases from the clinical practice of S.G.) highlight presentations for which different “wake-up calls” may be most effective.

### Vignette #1: Contamination OCD

Patient A was afraid of contracting a bloodborne pathogen such as Hepatitis B/C or AIDS through contact with “microscopic blood particles” in his environment. Patient A used barriers when he needed to touch something in public if he could not wash his hands immediately after. He wore long pants/shirts year-round, even during the summertime and despite overheating, to reduce risk of exposure. If something outside of his home touched his skin, Patient A would wash the area for 30 minutes. He constantly “tracked” what others touched to determine if any contaminated items had come in contact with things he would need to touch. He required his parents to buy Clorox wipes so that he could sanitize his phone and bag when he got home. He did not allow family members in his room for fear they might contaminate his “only safe space.” His OCD impaired his ability to attend school consistently and resulted in interpersonal difficulties with his friends and family.

### Treatment

Patient A found it difficult at first to remember to challenge his OCD when he started washing, especially when he was not in the clinic. To help cue Patient A to his exposure goals, Patient A and his therapist created visual cue cards using his favorite memes to catch his eye and included captions that reminded him of his exposure challenges (reducing washing time/steps and resisting tracking others). Patient A placed these cards on his bathroom mirror at eye level so he could not miss them, and on his desk next to his clock and computer. These cues helped Patient A be more consistent with his exposures and increase response prevention by redirecting his attention towards his exposure tasks.

### Vignette #2: Scrupulosity OCD

Patient B’s OCD focused on religious themes, and she spent nearly all day engaged in mental compulsions. She was afraid of accidentally practicing another religion and believed this would result in her eternal damnation. She repeated prayers and phrases both in her head and under her breath and felt compelled to repeat them until she recited them perfectly. Patient B described her obsessions and compulsions as automatic and uncontrollable. She stated, “sometimes I don’t even realize I am doing it until someone asks me what I am whispering. It is like they are on a never-ending loop. I never feel like I have more than a few seconds at a time without having to do one because a thought comes into my mind.” Patient B’s intrusions were nearly constant. This, in combination with the automatic nature of her compulsions, made exposure therapy particularly difficult for her to engage in.

### Treatment

For Patient B, building awareness of when she experienced obsessions and compulsions was the initial target of treatment. To help her build this awareness, Patient B’s therapist asked her to practice standing up out of her chair and saying a coping statement out loud (ex. “Maybe [that’s true/will happen], maybe not. Either way I am not going to do anything. Let’s see what happens.”) whenever she noticed an obsession. This reminded Patient B to start practicing a new pattern of thinking and acted as a competing response, because she was unable to recite prayers while saying her coping statement out loud. The purpose of standing up was to help Patient B break her obsessive-thought loop; it required several purposeful actions on Patient B’s part which helped her separate from her thoughts and focus on something external (the feeling of movement, seeing her therapist stand up with her each time) and played a function of externally orienting cue (a “wake-up call”). Patient B practiced this intervention for 10-minute intervals multiple times a day. She described her thoughts becoming “less sticky” with repeated practice. Once she could resist compulsions more readily, she was able to start new exposures that relied less on competing responses or grounding techniques and more on mindfully observing her experience.

## Discussion

An executive overload model of OCD (Abramovitch et al., 2012) suggests that internal preoccupation with obsessive-compulsive thoughts prevent individuals with OCD from fully engaging with external stimuli, contributing to difficulties with externally oriented executive functioning. Our behavioral data are consistent with these predictions and suggest that externally orienting cues (“wake-up calls”) may facilitate engagement with external stimuli, improve the efficacy of executive attention, and reduce difficulties with executive function. These effects are distinct for individuals with different OC themes. For instance, our results indicate that individuals with *int-OCs* (taboo, religious, or aggressive obsessions or mental compulsions) have more difficulty disengaging from internally oriented obsessive-compulsive loops and that performance on cognitive control tasks improves the most in individuals with *cont-OCs* (washing symptoms).

One of the reasons why some individuals do not respond to ERP-based interventions is their limited ability to adequately engage in the treatment (VanDyke & Pollard, 2005). In individuals with *int-OCs*, such engagement can be affected by reluctance to share the content of their taboo thoughts with the therapist, reduced awareness of their mental compulsions, and significant difficulty separating from internal obsessive thought loops and engaging with outside world. Our results indicate that individuals with *int-OCs* may also have difficulty attending to external stimuli during response prevention interventions. In the two clinical vignettes described above, a “wake-up call” was utilized to help patients separate from their internal world and attend to their external experience during response prevention exercises. For Patient A, a gentle visual reminder was all that was needed. For Patient B, a more complex and involved cue was needed to help pull her out of her thoughts and ground her in the moment. Both examples show how “wake-up calls” can be incorporated into ERP treatment to boost response prevention, which is the most crucial mechanism of change in ERP. Since not all patients will respond to such cues equally, the cue must be customized based off an individual’s ability to attend to external stimuli when prompted.

Note that “wake-up call” based interventions align well with the intuition that led to the development of mindfulness-based ERP (mb-ERP) approaches (Schoenberg et al., 2014; Strauss et al., 2015). Mindfulness can be defined as “paying attention in a particular way: on purpose, in the present moment, and nonjudgmentally” (Zinn, 1994). In relation to OCD, mindfulness trainings have been proposed to facilitate less reactivity to internal experience (Wilkinson-Tough et al., 2010) and to help patients to engage in the present to notice urges toward unhelpful, compulsive behaviors (e.g. hand washing or checking) and to engage instead in alternative, more helpful behavioral choices (Hale et al., 2013). However, there is an important difference between the stance of Mb-ERP and the idea of a “wake-up call” suggested by our results. Mb-ERP invites patients to attend to internal distressing thoughts and experiences nonjudgmentally and with acceptance, to facilitate fear reduction through safety learning and thus reduce the need for compensatory compulsions. In contrast, “wake-up calls” may help patients break away from involuntary mental activity (automatic thought patterns or mental compulsions) and attend to the external environment. Such external reorientation may reduce the risk of automatic compulsions and improve cognitive control over voluntary compulsions, including during response prevention exercises. Since difficulties with executive function have been linked to reduced quality of life in affected individuals (Berna Binnur et al., 2005; Subramaniam et al., 2013), future studies may examine a possibility that including “wake-up calls” in daily routines may also lead to greater improvements in overall functioning while living with OCs, outside of ERP. We call for future exploratory clinical trials to examine impacts of such interventions on both severity of OCs and on the overall quality of life, especially in ERP refractory patients.

The ANT-R task examines how improved readiness to respond to external stimuli by externally orienting cues impacts performance on two cognitive control tasks (Simon and Flanker). Incorporating similar “wake-up calls” into other broadly used tests of executive function in OCD (Snyder et al., 2015) may help to examine generalizability of our results across various domains of executive functions. Furthermore, it may help to examine whether internal preoccupation is a driving or just a contributing factor to the broad difficulties with executive functioning seen in OCD.

Examining behavioral tasks in combination with neuroimaging data may also help to evaluate the relative contribution of altered functioning in the cortico-striatal circuits (Nakao, Okada, & Kanba, 2014) and altered coordination among DMN, CEN, and SN (Posner et al., 2017) to difficulties with executive functioning in OCD. For instance, future neurobiological studies should explicitly test whether “wake-up calls” have an impact on the ability of SN to modulate DMN↔CEN switch in response to external stimuli. These studies also need to test whether this impact is stronger in individuals with *cont-OCs* and weaker in individuals with *int-OCs*. This line of research may lead to more nuanced versions of the DMN-ih.

Note that our study participants were not receiving any medications. Thus, our results do not speak to how “wake-up calls” might interact with medication effects. Follow up cross-sectional and longitudinal studies are needed to evaluate such effects. Another important limitation is our reliance on self-report measures of symptom severity that are subject to reporting biases (recall that some participants appeared to minimize their OCs).

Our exploratory analyses did not reveal any gender differences, but we did not control for the estradiol level, which has been shown to be associated with severity of OCs (Vulink et al., 2006) and more so with severity of *cont-OCs* (Labad et al., 2008).

Overall, our study is an example of how a specific phenomenon (e.g., an impact of preoccupation with OCs on externally oriented functioning) can be examined by integrating behavioral and clinical evidence; it also builds a roadmap for future incorporation of neurobiological data. To date, clinical investigations have accumulated significant amount of data at different level of analyses. Integrating such evidence to test and refine existing and generate new hypotheses can both strengthen empirical evidence and provide opportunities for more nuanced understanding of pathophysiology, which ultimately will lead to more effective treatments.

## Data Availability

All data produced in the present study are available upon reasonable request to the authors.

## ^1^Abbreviations

*OCD*: Obsessive-compulsive disorder
*OCs*: Obsessive-compulsive themes
*int-Ocs*: Obsessive-compulsive themes related to sexual, religious, unacceptable/taboo, and aggressive thoughts and/or involving mental compulsions
*cont-OCs*: Obsessive-compulsive themes related to contamination concerns
DMN: Default Mode Network
SN: Salience Network
CEN: Central Executive Network
DMN-ih: Default Mode Network interference hypothesis
ANT-R: Revised Attention Network Test
CBT: Cognitive Behavioral Therapy
ERP: Exposure and Response Prevention
ADHD: Attention Deficit Hyperactivity Disorder

## Acknowledgements

We thank Stephen A. Kichuk, Eileen Billingslea, and Ashton Megli for their support in subject recruitment and data collection and David Tolin for his insightful comments on earlier versions of the manuscript.

## Data statement

Data is available upon request.

