## Appendix for "Does Internal Preoccupation with Obsessive-Compulsive Themes Affect Externally Oriented Functioning in OCD?: Behavioral Results and Clinical Cases"

for

### Appendix A

*Sample Description*

**Table A.1**

*Descriptive statistics for the full sample (OCD N = 43, HC N = 54)*

| | All participants | OCD | | HC | | test stats | p |
|---|---|---|---|---|---|---|---|
| | mean (SD) | range | mean (SD) | range | mean (SD) | | |
| Male, n (%) | 38 (39) | 13 (34) | | 25 (46) | | $X^2$ = 2.6 | 0.11 |
| Age, N = 97 | 32.1 (11.8) | 18 - 63 | 34.3 (12.5) | 18 - 59 | 30.4 (11.0) | F = 2.7 | 0.10 |
| IQ, N = 96 | 110 (14) | 82 - 130 | 105 (12) | 77 - 132 | 113 (14) | F = 9.9 | 0.002 |
| Indecisiveness, N = 87 | 39.8 (10.9) | 15 - 69 | 42.5 (13.7) | 17 - 57 | 38.0 (8.3) | F = 3.6 | 0.06 |
| Y-BOCS total, N = 43 | - | 17 - 35 | 25.1 (4.3) | - | - | - | - |
| Y-BOCS obsessions | - | 0 - 17 | 12.0 (2.9) | - | - | - | - |
| Y-BOCS compulsions | - | 9 - 19 | 13.1 (2.3) | - | - | - | - |
| BDI-II, N = 95 | 6.6 (8.5) | 0 - 36 | 10.9 (10.1) | 0 - 20 | 3.1 (4.8) | M-W U = 1668 | <0.001 |
| SHAPS, N = 82 | 0.9 (2.1) | 0 - 8 | 0.7 (1.8) | 0 - 11 | 1.0 (2.3) | M-W U = 1780 | 0.90 |
| ASRS, N = 87 | 5.1 (4.8) | 0 - 18 | 6.5 (5.7) | 0 - 13 | 4.2 (3.8) | M-W U = 1102 | 0.09 |
| Alerting | 42.5 (31.9) | -40.6 - 106.4 | 37.4 (29.9) | -20.7 - 132.7 | 46.7 (33.1) | F = 2 | 0.16 |
| Executive, Simon | -3.5 (20.2) | -45.1 - 51.4 | -1.8 (19.1) | -43.2 - 49.6 | -4.9 (21.2) | F = 0.6 | 0.46 |
| Executive, Flanker | 141.7 (53.3) | 37.6 - 334.7 | 136.9 (57.1) | 39.3 - 309.9 | 145.4 (50.4) | M-W U = 1055 | 0.44 |
| Alerting x Simon | 10.9 (46.7) | -56.1 - 141.8 | 27.0 (44.5) | -109.4 - 79.0 | -1.9 (44.9) | F = 10 | 0.002 |
| Alerting x Flanker | 11.5 (51.5) | -78.0 - 145.1 | 14.3 (48.4) | -188.8 - 121.4 | 9.3 (54.2) | F = 0.2 | 0.64 |

*Note.* IQ - the Kaufman Brief Intelligence Test (Kaufman & Kaufman, 2004); Indecisiveness - Indecisiveness Scale (Frost & Shows, 1993); BDI - Beck Depression Inventory II (Beck et al., 1996); SHAPS - Snaith–Hamilton Pleasure Scale (Snaith et al., 1995); ASRS - Adult ADHD Self-Report Scale (Adler et al., 2006); Alerting, Executive Simon, Executive Flanker, Alerting x Simon, Alerting x Flanker – behavior-based measures from the Attentional Networks Test Revised (ANT-R). We tested the distribution of each variable of interest for normality (using the Shapiro Wilks test); we employed Mann-Whitney U test (M-W U stat) for measures that violated the assumption of normality, and 1 x 2 ANOVA (F-stat) for measures that were distributed near normally.

**Figure A.1**

*Distributions of race did not differ across diagnostic groups*

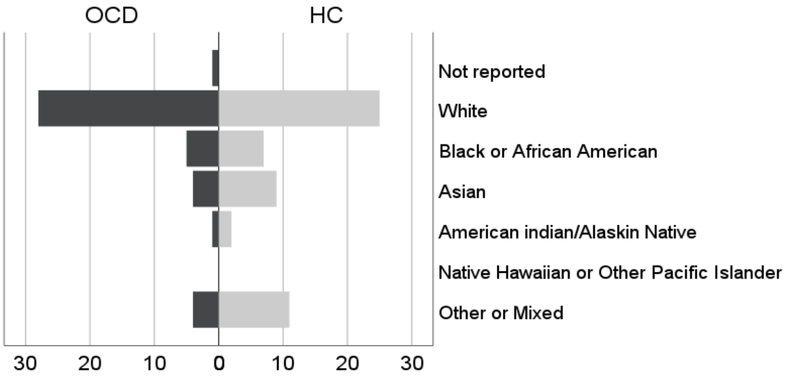


*Note:* $X^2 = 5.9$ $p = 0.32$

**Figure A.2**

*Distributions of education level did not differ across diagnostic groups*

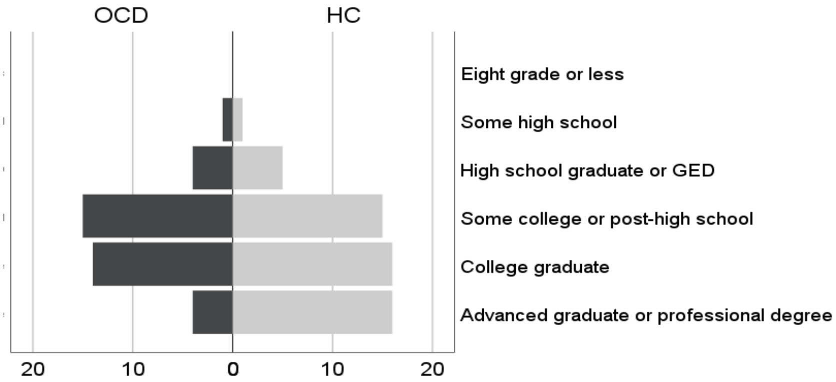


*Note:* $X^2 = 5.1$ $p = 0.28$

**Figure A.3**

*Individuals with OCD were less likely to belong to higher income groups than were healthy individuals*

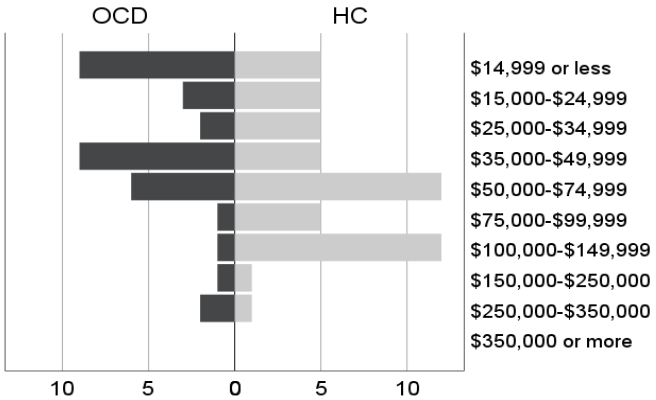


*Note:* $X^2 = 15.6$ $p = 0.048$

#### Appendix B

*Bivariate correlations among variables of interest*

**Table B.1**

*Spearman's correlations in OCI-R sample: potentially confounding variables*

| | | Alerting | Alerting x Simon | Alerting x Flanker | OCI-R | Washing | Obsessing | Hoarding | Ordering | Checking | Neutralizing |
|---|---|---|---|---|---|---|---|---|---|---|---|
| Age | ρ | 0.02 | -0.03 | -0.11 | 0.05 | 0.09 | -0.05 | -0.18 | -0.007 | 0.08 | 0.12 |
| | p | 0.90 | 0.79 | 0.34 | 0.69 | 0.44 | 0.64 | 0.11 | 0.95 | 0.47 | 0.30 |
| IQ | ρ | -0.09 | 0.05 | 0.05 | -0.06 | -.22* | 0.01 | .26* | 0.07 | -0.18 [a] | 0.02 |
| | p | 0.40 | 0.67 | 0.65 | 0.62 | 0.04 | 0.95 | 0.02 | 0.51 | 0.10 | 0.88 |
| BDI-II | ρ | -0.20 | -0.07 | -0.05 | 0.46*** | 0.28** | 0.57** | 0.34*** | 0.29** | 0.31** | 0.38*** |
| | p | 0.07[a] | 0.55 | 0.67 | < 0.001 | 0.01 | < 0.001 | 0.001 | 0.007 | 0.004 | < 0.001 |
| ARSR | ρ | -0.17 | -0.06 | -0.003 | 0.33** | 0.18 [a] | 0.48** | 0.37** | 0.31** | 0.12 | 0.28** |
| | p | 0.12 | 0.62 | 0.98 | 0.002 | 0.10 | < 0.001 | 0.001 | 0.005 | 0.29 | 0.01 |

*Note.* Significance levels *** - $p < 0.001$; ** - $p < 0.01$; * - $p < 0.05$ level; [a] - $p < 0.1$. <u>Not included in the table:</u> BDI-II did not significantly correlate with age and IQ ( $p > 0.34$); ASRS negatively correlated with age ($\rho = -0.22$ $p = 0.04$) and positively at a trend level with IQ ($\rho = 0.19$ $p = 0.09$); BDI-II and ASRS positively correlated ($\rho = 0.53$ $p < 0.001$).

**Table B.2**

*Spearman's correlations in OCI-R sample: OCI-R subscales standardized residuals*

| | | OCI-R, stdres | Washing, stdres | Obsessing, stdres | Hoarding, stdres | Ordering, stdres | Checking, stdres | Neutralizing, stdres |
|---|---|---|---|---|---|---|---|---|
| age | ρ | -0.06 | -0.10 | -0.06 | 0.06 | -0.06 | -0.07 | -0.06 |
| | p | 0.57 | 0.39 | 0.61 | 0.58 | 0.60 | 0.54 | 0.59 |
| IQ | ρ | 0.05 | 0.17 | 0.001 | 0.01 | 0.05 | 0.13 | -0.02 |
| | p | 0.67 | 0.13 | 0.99 | 0.95 | 0.66 | 0.23 | 0.84 |
| BDI-II | ρ | 0.002 | -0.08 | -0.06 | -0.07 | -0.02 | -0.07 | -0.22* |
| | p | 0.99 | 0.49 | 0.61 | 0.52 | 0.87 | 0.56 | 0.04 |
| ASRS | ρ | -0.01 | -0.04 | -0.07 | -0.09 | 0.003 | 0.03 | -0.10 |
| | p | 0.92 | 0.71 | 0.51 | 0.41 | 0.98 | 0.80 | 0.38 |
| OCI-R | ρ | **0.81**** | 0.52** | 0.50** | 0.37** | 0.71** | 0.63** | 0.34** |
| | p | **< 0.001** | < 0.001 | < 0.001 | 0.001 | < 0.001 | < 0.001 | 0.002 |
| Washing | ρ | 0.64** | **0.78**** | 0.45** | 0.09 | 0.53** | 0.47** | 0.10 |
| | p | < 0.001 | **< 0.001** | < 0.001 | 0.42 | < 0.001 | < 0.001 | 0.37 |
| Obsessing | ρ | 0.44** | 0.26* | **0.64**** | 0.01 | 0.31** | 0.38** | 0.11 |
| | p | < 0.001 | 0.02 | **< 0.001** | 0.92 | 0.005 | < 0.001 | 0.32 |
| Hoarding | ρ | 0.45** | 0.10 | 0.11 | **0.79**** | 0.31** | 0.35** | 0.22* |
| | p | < 0.001 | 0.35 | 0.33 | **< 0.001** | 0.005 | 0.001 | 0.04 |
| Ordering | ρ | 0.75** | 0.48** | 0.43** | 0.29** | **0.90**** | 0.52** | 0.29** |
| | p | < 0.001 | < 0.001 | < 0.001 | 0.01 | **< 0.001** | < 0.001 | 0.009 |
| Checking | ρ | 0.76** | 0.42** | 0.46** | 0.31** | 0.54** | **0.83**** | 0.32** |
| | p | < 0.001 | < 0.001 | < 0.001 | 0.004 | < 0.001 | **< 0.001** | 0.004 |
| Neutralizing | ρ | 0.46** | 0.09 | 0.23* | 0.25* | 0.32** | 0.34** | **0.69*** |
| | p | < 0.001 | 0.413 | 0.03 | 0.02 | 0.003 | 0.002 | **< 0.001** |
| Alerting | ρ | 0.001 | 0.18 | -0.05 | -0.07 | 0.09 | -0.04 | -0.14 |
| | p | 0.99 | 0.11 | 0.64 | 0.52 | 0.41 | 0.72 | 0.20 |
| Alerting x | ρ | 0.36** | 0.32** | 0.25* | -0.02 | 0.31** | 0.31** | 0.21[a] |
| Simon | p | 0.001 | 0.003 | 0.03 | 0.84 | 0.004 | 0.005 | 0.06 |
| Alerting x | ρ | 0.15 | 0.35** | -0.06 | 0.03 | 0.07 | 0.15 | -0.10 |
| Flanker | p | 0.19 | 0.001 | 0.60 | 0.78 | 0.55 | 0.19 | 0.36 |

*Note.* Significance levels *** - p < 0.001; ** - p < 0.01; * - p < 0.05 level; [a] - p < 0.1.; stdres – standardized residuals; in **bold** - correlations between an OCI-R (sub)scale and the corresponding standardized residual.

#### Appendix C

*Descriptive analyses of measures relevant for testable hypotheses*

**Figure C.1**

*Proportion of trails with correct responses, by OCD diagnosis*

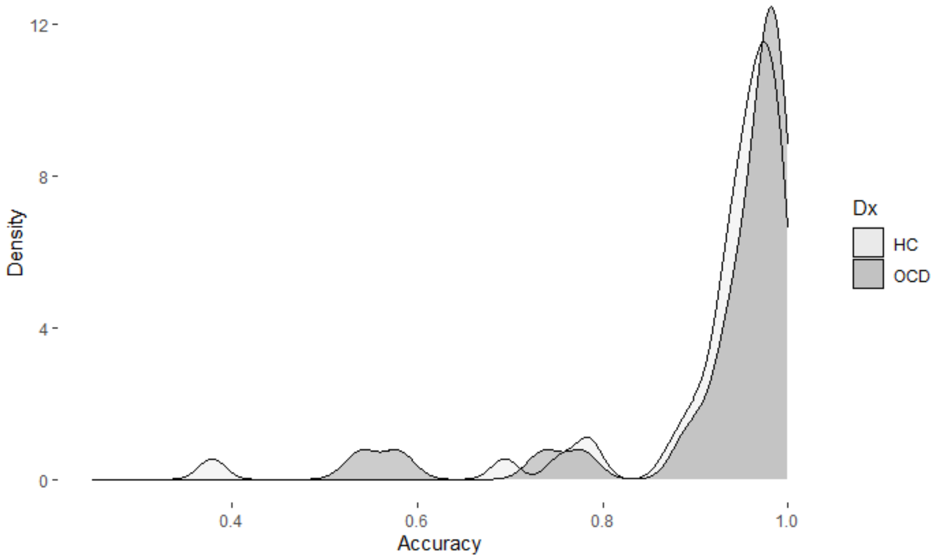


*Note.* Consistent with prior studies, most participants responded correctly on vast majority of trials.

**Figure C.2**

*Variables represented on the horizontal axis of Figure 1 (main text)*

**C.2.1**

*Distribution of severity of OCs, by OCD diagnosis; OCI-R subsample*

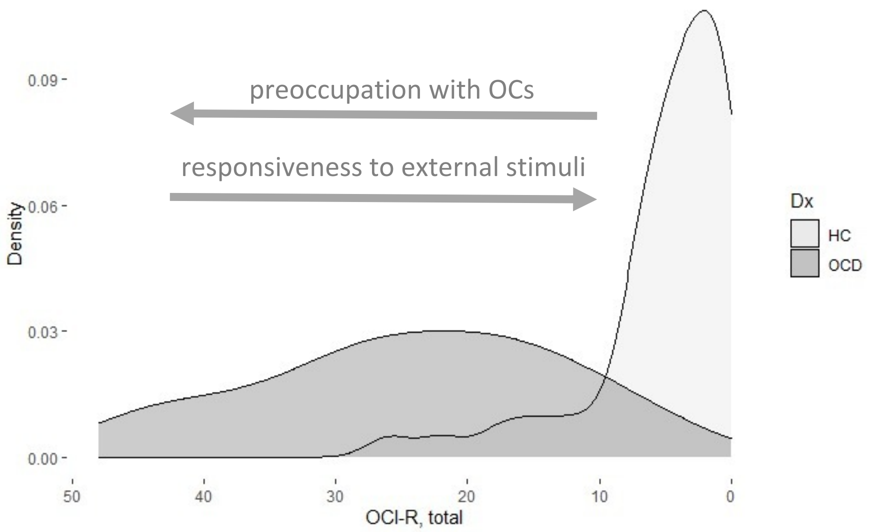


*Note.* OCs distributions were broad and overlapping across two diagnostic groups.

**C.2.2**

*Boxplots of RT $_{no\ cue}$, RT $_{double\ cue}$, and Alerting benefits (RT $_{no\ cue}$ - RT $_{double\ cue}$), by OCD diagnosis; matched subsample*

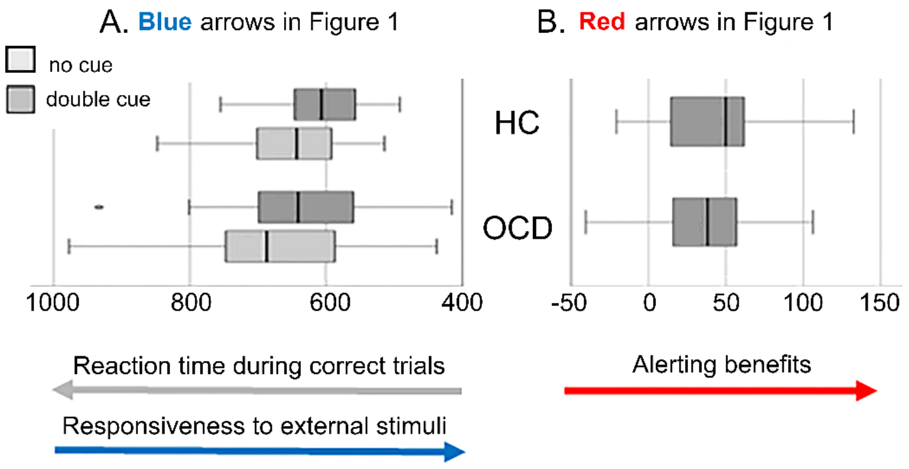


*Note.* OCD tended to respond slower than HC in no cue trials (**blue** arrows in Figure 1). RT improvements from no cue condition to double cue condition were qualitatively smaller in OCD than in HC (**red** arrows in Figure 1); black lines indicate medians.

**Figure C.2**

*Variables represented on the vertical axis of Figure 1 (main text)*

**C.2.1**

*Costs of processing Simon conflict and effects of alerting benefits on Simon costs, by OCD diagnosis*

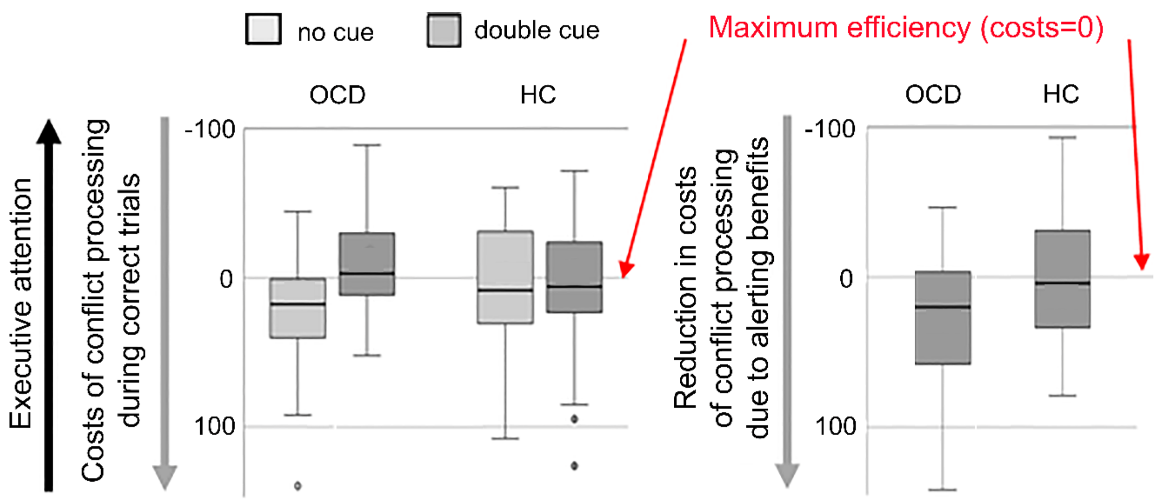


**C.2.2**

*Costs of processing Flanker conflict and effects of alerting benefits on Flanker costs, by OCD diagnosis*

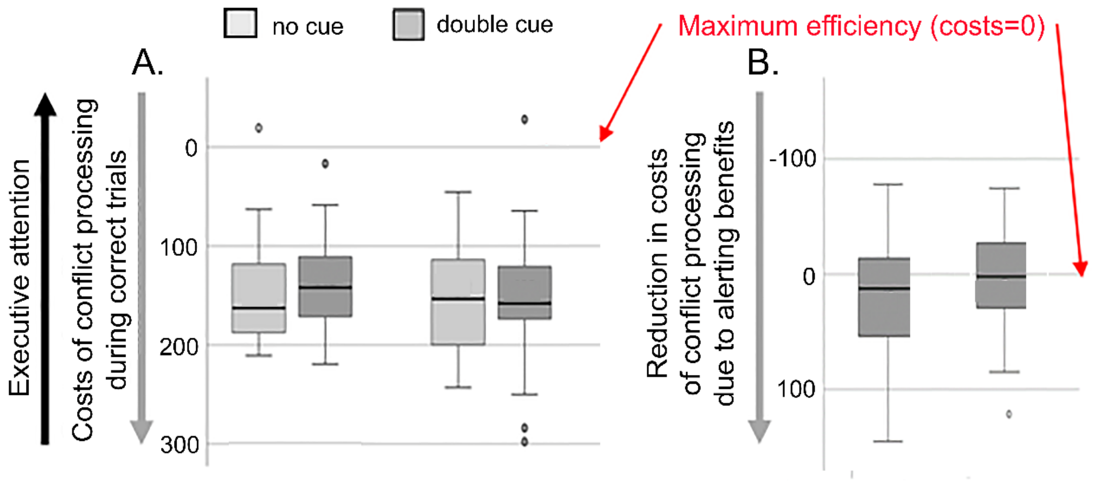


*Note*. Conflict processing costs in OCD were qualitatively higher than in HC during no cue trials (lower efficiency of executive attention in OCD; left panels); reduction in conflict processing costs from no cue condition to double cue condition were qualitatively larger in OCD than in HC (higher alerting x executive interactions; right panel); black lines indicate medians.

#### Appendix D

*Testing study hypotheses in OCD subsample, N = 32*

**Table D.1**

*Testing H1a: univariate ANCOVA with dependent variable Alerting*

| Predictor | effect | | ANCOVA | | parameter estimates | |
|---|---|---|---|---|---|---|
| | direction | size, $\eta^2$ | F | p | β | SE |
| Constant | + | 0.79*** | 76.7 | < 0.001 | 38.36 | 4.38 |
| age, std | - | < 0.001 | 0.005 | 0.94 | -0.35 | 4.69 |
| IQ, std | + | 0.005 | 0.10 | 0.75 | 1.46 | 4.59 |
| ASRS, std | - | 0.003 | 0.06 | 0.81 | 1.61 | 6.72 |
| BDI, std | + | 0.03 | 0.69 | 0.42 | -5.61 | 6.75 |
| Washing, stdres | + | 0.13[a] | 3.05 | 0.095 | 11.28 | 6.46 |
| Obsessing, stdres | - | 0.22* | 6.07 | 0.02 | -15.14 | 6.15 |
| Hoarding, stdres | - | 0.07 | 1.56 | 0.23 | -7.41 | 5.93 |
| Ordering, stdres | + | 0.12[a] | 2.95 | 0.10 | 12.16 | 7.08 |
| Checking, stdres | + | < 0.001 | < 0.001 | 0.98 | 0.16 | 7.15 |
| Neutralizing, stdres | - | 0.16[a] | 3.89 | 0.06 | -10.70 | 5.43 |

*Note.* Significance levels *** - p < 0.001; ** - p < 0.01; * - p < 0.05 level; [a] - p < 0.1. Effect directions + - positive, - - negative. Effect sizes $\eta^2$ = 0.01 - small effect, $\eta^2$ = 0.06 - medium effect, $\eta^2$ = 0.14 - large effect. Covariates std – standardized, stdres – standardized residuals.

**Table D.2**

*Testing H2a: repeated measures ANCOVA with dependent variable Alerting x Executive, and repeated measures type of conflict (Simon/Flanker)*

| Predictors | Main effects | | | | Predictor x conflict interactions | | | |
|---|---|---|---|---|---|---|---|---|
| | effect | | ANCOVA | | effect | | ANCOVA | |
| | direction | size, $\eta^2$ | F | p | direction | size, $\eta^2$ | F | p |
| Intercept | + | 0.28 | 0.96 | 0.34 | S > F | 0.05 | 0.96 | 0.34 |
| age, std | - | 0.005 | 0.02 | 0.90 | S < F | 0.001 | 0.02 | 0.90 |
| IQ, std | n.c. | 0.04 | 2.19 | 0.15 | n.c. | 0.10 | 2.19 | 0.15 |
| ASRS, std | n.c. | 0.05 | 2.70 | 0.12 | n.c. | 0.12 | 2.70 | 0.12 |
| BDI, std | n.c. | < 0.001 | 0.64 | 0.43 | n.c. | 0.03 | 0.64 | 0.43 |
| Alerting, std | n.c. | < 0.001 | 0.33 | 0.57 | n.c. | 0.02 | 0.33 | 0.57 |
| Washing, stdres | + | 0.07 | 1.26 | 0.27 | S < F | 0.06 | 1.26 | 0.27 |
| Obsessing, stdres | n.c. | 0.02 | 2.34 | 0.14 | n.c. | 0.11 | 2.34 | 0.14 |
| Hoarding, stdres | + | 0.001 | 0.02 | 0.90 | S < F | 0.001 | 0.02 | 0.90 |
| Ordering, stdres | + | 0.02 | 0.52 | 0.48 | S > F | 0.03 | 0.52 | 0.48 |
| Checking, stdres | n.c. | < 0.001 | 2.83 | 0.11 | n.c. | 0.13 | 2.83 | 0.11 |
| Neutralizing, stdres | n.c. | 0.03 | 2.73 | 0.11 | n.c. | 0.12 | 2.73 | 0.11 |

*Note*. Main effect directions + - positive, - - negative; difference of effects across types of conflicts S>F – effect is stronger under Simon conflict, S<F – effect is stronger under Flanker conflict, n.c. – no consistent pattern as effects change direction across conflicts. Effect sizes $\eta^2$ = 0.01 - small effect, $\eta^2$ = 0.06 - medium effect, $\eta^2$ = 0.14 - large effect. Covariates std – standardized, stdres – standardized residuals. The effect of washing is of medium-to-large size but did not reach statistical significance.

**Appendix E**

*Testing study hypotheses without controlling for confounding effects of depression and ADHD*

**Table E.1**

*Testing H1: univariate ANCOVA with dependent variable Alerting*

| Predictor | effect | | ANCOVA | | parameter estimates | |
|---|---|---|---|---|---|---|
| | direction | size, $\eta^2$ | F | p | β | SE |
| Constant | + | 0.99 [a] | 99.79 | 0.07 | 37.32 | 5.88 |
| age, std | + | 0.002 | 0.17 | 0.68 | 1.54 | 3.69 |
| IQ, std | - | 0.006 | 0.46 | 0.50 | -2.69 | 3.98 |
| Dx | + | 0.02 | 1.20 | 0.28 | 8.36 | 7.62 |

*Note.* Significance levels [a] - $p < 0.1$. Main effect directions + - positive, - - negative; difference of effects across types of conflicts S>F – effect is stronger under Simon conflict, S < F – effect is stronger under Flanker conflict. Effect sizes $\eta^2 = 0.01$ - small effect, $\eta^2 = 0.06$ - medium effect, $\eta^2 = 0.14$ - large effect. Covariates std – standardized.

**Table E.2**

*Testing H1a: univariate ANCOVA with dependent variable Alerting*

| Predictor | effect | | ANCOVA | | parameter estimates | |
|---|---|---|---|---|---|---|
| | direction | size, $\eta^2$ | F | p | β | SE |
| Constant | + | 0.69 | 164.62 | 0.00 | 42.76 | 3.33 |
| age, std | + | 0.00 | 0.19 | 0.66 | 1.53 | 3.48 |
| IQ, std | + | 0.00 | 0.00 | 0.95 | 0.23 | 3.81 |
| Washing, std | + | 0.01 | 0.98 | 0.32 | 5.09 | 5.13 |
| Obsessing, std | - | 0.04[a] | 3.47 | 0.07 | -8.93 | 4.79 |
| Hoarding, std | - | 0.02 | 1.81 | 0.18 | -5.49 | 4.08 |
| Ordering, std | + | 0.05[a] | 3.75 | 0.06 | 11.19 | 5.78 |
| Checking, std | - | 0.00 | 0.24 | 0.62 | -2.52 | 5.12 |
| Neutralizing, std | - | 0.05* | 4.26 | 0.04 | -8.71 | 4.22 |

*Note.* Significance levels * - $p < 0.05$ level; [a] - $p < 0.1$. Effect directions + - positive, - - negative. Effect sizes $\eta^2 = 0.01$ - small effect, $\eta^2 = 0.06$ - medium effect, $\eta^2 = 0.14$ - large effect. Covariates std – standardized.

**Table E.3**

*Testing H2: repeated measures ANCOVA with dependent variable Alerting x Executive, and repeated measures type of conflict (Simon/Flanker)*

| Predictors | Main effects | | | | Predictor x conflict interactions | | | |
|---|---|---|---|---|---|---|---|---|
| | effect | | ANCOVA | | effect | | ANCOVA | |
| | direction | size, $\eta^2$ | F | p | direction | size, $\eta^2$ | F | p |
| Intercept | + | 0.07** | 6.28 | 0.01 | S > F | 0.01 | 0.81 | 0.37 |
| age, std | - | 0.01 | 0.72 | 0.40 | S < F | <0.001 | 0.01 | 0.93 |
| IQ, std | + | 0.003 | 0.26 | 0.61 | S > F | 0.003 | 0.23 | 0.63 |
| Alerting, std | + | 0.06* | 5.57 | 0.02 | S < F | 0.07* | 5.95 | 0.02 |
| Dx | + | 0.12** | 10.55 | 0.002 | S > F | 0.01 | 0.71 | 0.40 |

*Note.* Significance levels ** - $p < 0.01$; * - $p < 0.05$ level. Main effect directions + - positive, - - negative; difference of effects across types of conflicts S>F – effect is stronger under Simon conflict, S < F – effect is stronger under Flanker conflict. Effect sizes $\eta^2 = 0.01$ - small effect, $\eta^2 = 0.06$ - medium effect, $\eta^2 = 0.14$ - large effect. Covariates std – standardized.

**Table E.4**

*Testing H2a: repeated measures ANCOVA with dependent variable Alerting x Executive, and repeated measures type of conflict (Simon/Flanker)*

| | Main effects | | | | Predictor x conflict interactions | | | |
|---|---|---|---|---|---|---|---|---|
| Predictors | effect | | ANCOVA | | effect | | ANCOVA | |
| | direction | size, $\eta^2$ | F | p | direction | size, $\eta^2$ | F | p |
| Intercept | + | 0.08** | 6.57 | 0.01 | S < F | 0.001 | 0.08 | 0.78 |
| age, std | - | 0.02 | 1.12 | 0.29 | S < F | 0.003 | 0.23 | 0.63 |
| IQ, std | + | 0.06* | 4.27 | 0.04 | S < F | 0.01 | 0.63 | 0.43 |
| Alerting, std | n.c. | 0.02 | 1.14 | 0.29 | n.c. | 0.09** | 7.55 | 0.01 |
| Washing, std | + | 0.12** | 9.80 | 0.003 | S < F | 0.003 | 0.21 | 0.65 |
| Obsessing, std | - | 0.06* | 4.32 | 0.04 | S < F | 0.001 | 0.06 | 0.81 |
| Hoarding, std | n.c. | 0.006 | 0.41 | 0.53 | n.c. | 0.05a | 3.46 | 0.07 |
| Ordering, std | n.c. | 0.004 | 0.26 | 0.61 | n.c. | 0.03 | 1.89 | 0.17 |
| Checking, std | + | 0.04[a] | 3.05 | 0.08 | S < F | 0.01 | 0.78 | 0.38 |
| Neutralizing, std | n.c. | <0.001 | 0.00 | 0.99 | n.c. | 0.03 | 2.38 | 0.13 |

*Note.* Significance levels ** - p < 0.01; * - p < 0.05 level; [a] - p < 0.1. Main effect directions + - positive, - - negative; difference of effects across types of conflicts S>F – effect is stronger under Simon conflict, S < F – effect is stronger under Flanker conflict, n.c. – no consistent pattern as effects change direction across conflicts. Effect sizes $\eta^2$ = 0.01 - small effect, $\eta^2$ = 0.06 - medium effect, $\eta^2$ = 0.14 - large effect. Covariates std – standardized.
